# Effectiveness of using implementation frameworks to facilitate the implementation of a stroke management guideline in the Traditional Chinese Medicine hospitals in China: protocol for a factorial randomized controlled trial

**DOI:** 10.1101/2024.06.20.24309138

**Authors:** Wenjun He, Yiyuan Cai, Chun Hao, Zhuo Chen, Yuning Shi, Pengfei Guo, Sensen Lv, Lanping Zhang, Qing Zhao, Lingrui Liu, Yefeng Cai, Dong (Roman)Xu

## Abstract

**Background:** Traditional Chinese medicine (TCM) is commonly used alongside western medicine for stroke management in China. However, there is significant variation in TCM practice, and the utilization of evidence-based clinical practice guidelines is inadequate.This study aims to evaluate the effectiveness of three popular frameworks (Consolidated Framework for Implementation Research, Theoretical Domains Framework, and Normalization Process Theory) in improving implementation outcomes for the integrated TCM and western medicine clinical practice guideline for stroke management.

**Methods:** The study will employ a hybrid type III design, utilizing a factorial randomized controlled approach. Implementation facilitators will be trained and randomly assigned to participating organizations. Forty-five TCM hospitals will be randomly assigned to one of eight experiment conditions, allowing for the evaluation of each framework’s main effect, two-way interactions, and three-way interaction. The outcomes will be assessed using the RE-AIM framework, including reach, effectiveness, adoption, implementation, and maintenance. Hierarchical logistic regression models will be used to test the hypotheses. Qualitative methods, such as interviews and focus groups, will provide contextual information, and a Cost-effectiveness analysis (CEA) will be conducted.

**Discussion:** This proposed hybrid type III study will integrate implementation science(frameworks), epidemiology (factorial design), and health economic approaches (CEA) to advance our empirical understanding of the strengths and weaknesses of implementation frameworks, implementing the TCM guideline promotion practices in hospitals and its health economic evaluation. The study answers the question that which framework combination works well in facilitating the implementation of clinical intervention. The results of the trial will offer valuable insights into how best to implement evidence-based practices (TCM guidelines) in hospitals under the consideration of economic evaluation.

## 1 Background

Adherence to credible and evidence-based clinical practice guidelines is a critical indicator of high-quality care. There has been a plethora of clinical practice guidelines for cardiovascular conditions worldwide. But in many countries, including China, guidelines are largely not implemented^1^. Stroke is the leading burden of disease in China. Inpatient stroke management has often featured a combined use of western and traditional Chinese medicine (TCM) in China. A cross-sectional survey involving 48 general hospitals across China showed that about one-third of patients with ischemic stroke received a wide range of proprietary Chinese medicine during their hospital care^2^. Another prospective observational study found that the use of TCM in new ischemic stroke patients was as high as 83.1%, even exceeding the use of antithrombotic drugs^3^. Furthermore, the most important clinical indication for acupuncture is the rehabilitation of post-stroke sequelae^4,5^. The clinical practice guideline to promote evidence-based practice in managing stroke with integrated western and traditional Chinese medicine has been developed through a rigorous evidence-based medicine approach^5^. But the resistance to using the guideline in clinical routines has remained high. Few studies in China have been conducted to develop and test the effectiveness of implementation strategies to improve the normalization of the guidelines into the practice routine among inpatient stroke management.

Theories, models, and frameworks (hereafter referred to as “frameworks”) are widely used in implementation research. They provide systematic guidance on determining the implementation process, identifying the determinants (barriers and facilitators) of implementing evidence-based practice, and evaluating the effect of the implementation strategy. The frameworks have been used to identify implementation determinants, inform data collection, enhance conceptual clarity, and guide implementation planning^6^. The reliance on the frameworks has become the hallmark of implementation research. Three popular frameworks - Consolidated Framework for Implementation Research (CFIR), the Normalization Process Theory (NPT), and the Theoretical Domain Frameworks (TDF) have received a staggering12845 citations in total^7–9^ (Web of Knowledge accessed Oct. 2022), indicating widespread use in research. However many frameworks assumed the validity of their effectiveness in improving implementation with insufficient validation evidence. Even fewer studies provided empirical evidence for their effectiveness. The debates continue regarding whether the use of the frameworks provided advantages over the use of human instinct^10^. There has also been little guidance on which frameworks work better in improving guideline implementation under certain contexts. Furthermore, frameworks can often be used in conjunction. So it’s not only necessary to investigate the use of a single framework but also important to understand the interaction among frameworks. Factorial design can be an efficient trial design to examine both the main effects and the interaction effects.

Moreover, in light of these theoretical frameworks, successful behavior change efforts in implementing evidence-based practices (EBP) occur not only in the inner setting where clients and practitioners reside but also in an outer setting, where the organization and social context reside^11^. Contextual factors and organizational constructs or processes are associated with the dissemination and adoption of EBPs in various settings^12,13^. In the present project, we propose to employ an innovative method, named Qualitative Comparative Analysis (QCA^14^), to identify the configurations (combinations) of the setting features, that when present in the practices, could at most contribute to the implementation outcomes. QCA, a method that originated in political science and sociology, has demonstrated its utility in public health evaluative science^15^.

In summary, this study will address two research questions: (1) whether implementation theories, models, and frameworks, alone and in conjunction, promote the implementation of a clinical practice guideline for stroke management in Chinese hospitals, and (2) what combination of contextual factors contributes most to the implementation of this stroke guideline. The former question will be investigated with an experimental design, while the latter will use the mixed method of qualitative comparative analysis.

## 2 Method

### 2.1 Study design

This manuscript adheres to the Standards for Reporting Implementation Studies (StaRI) Statement (Additional file 3)^16^.

This will be a hybrid type III study^17^ whereby we will primarily test the effect of the implementation strategy in the real-world setting while gathering information on the effectiveness of the clinical intervention (i.e., the use of the guideline in this study). Considering the relatively weak evidence base for TCM, it is necessary to collect health and clinical outcomes. However, in this hybrid type III trial, as all groups will be implementing the guideline (i.e., conducting the clinical intervention), we will not be able to compare the health outcomes between the groups implementing the guideline to the control groups not implementing it. Instead, the health and clinical outcomes will be evaluated in the factorial design described below.

The overall study will be based on a factorial cluster randomized controlled trial (RCT) design. The design will have three factors, each corresponding to the use of a framework of TDF, CFIR, or NPT. Each factor has two levels: in use and not in the use of a given framework. Thus, the complete use of those three factors (frameworks) will form 2×2×2=8 possible configurations or experiment conditions (Table 1). Those experiment conditions will be randomly assigned to the sampled participating organizations, with each participating organization subject to one experiment condition through random assignment. After participants have been exposed to a condition, the assessment measures of outcomes will be used to determine if any changes could be attributed to the experimental conditions. The factorial experiment does not have a fixed and single control group as in a conventional RCT. This complete 2×2×2 factorial design enables estimation of the main effect of each factor, three two-way interactions, and a three-way interaction^18^. The main effect of a factorial experiment is the effect between the two levels of a factor, collapsing over the levels of all remaining factors. For instance, the main effect of using NPT versus not using it in guiding the implementation of the stroke guideline is the difference in means of experiment condition (EC) 1-4 vs. EC 5-8, averaging over the levels of the other two factors TDF and CFIR. We select the factorial experiment mainly because of (1) the high efficiency of the design (with the same sample size, it offers much higher statistical power than a conventional RCT^19^), and (2) the possibility to examine interactions between the use of frameworks (e.g., we need to understand whether NPT works better when it is accompanied by the use of CFIR).

**Table 1.** Experiment conditions.

| Experiment Conditions | NPT | CFIR | TDF | Mean of Outcomes |
| --- | --- | --- | --- | --- |
| EC<sup>1</sup> 1 | -<sup>2</sup> | - | - | μ- - -<sup>3</sup> |
| EC 2 | - | - | + | μ- - + |
| EC 3 | - | + | - | μ- + - |
| EC 4 | - | + | + | μ- + + |
| EC 5 | + | - | - | $\mu$+ - - |
| EC 6 | + | - | + | $\mu$+ - + |
| EC 7 | + | + | - | $\mu + + -$ |
| EC 8 | + | + | + | $\mu + + +$ |

Note: <sup>1</sup>EC = experiment conditions; <sup>2</sup>“-” stands for “not having it” while “+” for “having it” in this experiment condition; <sup>3</sup> $\mu$ - - -: the mean of the outcome when none of the frameworks are used.

### 2.2 Procedures

#### 2.2.1 Selection of the frameworks

We have selected three implementation science frameworks (NPT, CFIR, and TDF) for this experiment. The selection of those three frameworks is based on the following criteria: (1) highly popular frameworks in implementation research^6^ (NPT, CFIR, and TDF have received 559, 6760, and 863 citations in peer-reviewed papers as of the year 2021), (2) potential applicability across the entire process of implementation (all three frameworks can, in theory, be used throughout the implementation process from the beginning to the end, despite their relative perceived strengths in a certain phase of the implementation), and (3) potential applicability of combined use with other frameworks.

#### 2.2.2 Experiment conditions

As aforementioned, those three frameworks establish eight experiment conditions ranging from the combined use of all three to the use of none and anything in between. The use of the framework will follow a standard procedure. The procedure will be developed through a modified Delphi process. Based on literature and empirical evidence, the research team will first develop a candidate procedural steps of the implementation procedure. Then a group of 10-15 implementation researchers who are otherwise not associated with the program will review and rate the procedure through several rounds of internet-based surveys. The Delphi rounds will stop when any of the following conditions are met: consensus reached (defined as Kendall’s W over 0.7), no significant difference between the expert opinions between two successive rounds (defined as not to reject the null hypothesis that there will be an equal number of changes in both directions in the target experts using McNemar chi-square tes^20^), or a maximum of four rounds reached^21^. Inspired by the NIATx Model^22^, the procedure as it stands now may have the following candidate steps:

1. Assemble an implementation team at each of the participating hospitals;
2. Identify the barriers and facilitators of implementing the stroke guideline;
3. 3 Develop and/or identify appropriate implementation techniques to address the barriers and enhance the facilitators;
4. Assign roles and tasks to the implementation team members to implement the selected techniques;
5. Monitor the implementation process and make necessary adaptations in implementation techniques;
6. Evaluate the effect of the implementation.

For each step, the team will be asked to consider the use of the frameworks when appropriate in guiding their investigation. The team has the flexibility in deciding how exactly they will conduct each step. For instance, for step two, they can use focus groups, interviews, nominal group techniques, or Delphi surveys to identify barriers and facilitators of implementing the guideline^23,24^, while using the selected frameworks to guide the content domains of the discussion. As another example, for step 5, they can use rapid cycle techniques such as the Plan-Do-Study-Act (PDSA) ^25^technique to gather rapid feedback and make necessary changes to the implementation plans.

During the implementation, each participating hospital will receive assistance from a pair of facilitators. Each facilitating pair will consist of a master’s degree student in health management and an undergraduate student in health-related sciences. All facilitators have already completed and passed the study of a 30-hour Introductory Course on Implementation Science, which covers the concept of implementation, implementation frameworks, implementation process, and unique research designs and methods (both qualitative and quantitative) in implementation research. The facilitators will be randomized into eight teams, corresponding to the eight experiment conditions (Table 1). The purpose of this randomization is to minimize the influence of the possible differences in implementation competence between the teams. Throughout the implementation, the facilitators will be instructed to use the selected frameworks in that experiment condition only. A Ph.D. student who is knowledgeable about implementation science will serve as the central adviser for all facilitators. The duration of the entire study was 1 year, and the implementation strategy will be stopped after 6 months of implementation, and the samples will be collected for a full year.

#### 2.2.3 Clinical intervention: TCM guideline

The clinical intervention to be implemented in this study is the Clinical Practice Guideline for the Management of Stroke with Integrated Traditional Chinese Medicine and Western Medicine. The guideline was developed by a multidisciplinary team of experts organized by Guangdong Provincial TCM Hospital (one of the largest academic TCM health centers in China) with methodological support from the GRADE China center at Lanzhou University. The development of this guideline followed the procedures and principles from the World Health Organization Handbook for Guideline Development, the Manual for the Development of Integrative Medicine Guidelines, as well as the Appraisal of Guidelines for Research and Evaluation, AGREE II^26–29^. The guideline^30^ gave 14 recommendations regarding the use of TCM in managing stroke, covering eight clinical questions: (1) “effect of TCM on neurological function in patients with ischemic stroke”, (2) secondary prevention of ischemic stroke and transient ischemic attack by TCM, (3) the effect of blood-activating herbs on patients with acute cerebral infarction thrombolysis, (4) effects of blood-activating herbs on patients transformed by cerebral infarction hemorrhage, (5) effects of blood-activating herbs on patients with hypertensive cerebral hemorrhage, (6) herbal treatment of stroke with impaired consciousness, (7) Chinese medicine treatment for post-stroke dysphagia, and (8) herbal treatment of post-stroke depression. The guideline, however, acknowledges that the overall evidence base for TCM is poor so that almost all recommendations were “suggested for consideration” rather than “mandatory”. Yet, this does not diminish the significance of the guideline as it substantially limits the use of numerous Chinese medicines to a few that have some evidence base.

### 2.3 Targeted sites and population

The study will engage four levels of participants related to stroke management in descending order: hospitals, clinical departments, physicians, and patients. The study will be conducted among the traditional Chinese medicine hospitals (TCM Hospitals) in Guangdong province. Despite their name, all those hospitals use both western medicine and traditional Chinese medicine to manage stroke. The practicing doctors also received training in western medicine and traditional Chinese medicine. It should be noted that TCM in China is not complementary medicine but in the mainstream. Targeting this group of hospitals is because, although hospitals practicing western medicine also use traditional Chinese medicine in their management of stroke to a large extent, those TCM hospitals more heavily use traditional Chinese medicine. Also, the clinicians with TCM specialty traditionally are less interested in basing their practice on evidence-based medicine. Guangdong is among the most economically developed provinces in China with a per capita GDP of 87899 RMB (ranked 7th among China’s 34 provinces) in 2020 and a life expectancy of 78.18 (ranked 8th) in 2019. Guangdong is also a stronghold for TCM use^31^. The province has developed a network of TCM hospitals for the management of brain conditions. Currently, with a total of 45 member hospitals, the network has included mostly county-level hospitals. China’s hospitals are classified into levels 1, 2, and 3 with the increasing sophistication of services. County hospitals are mainly level-two hospitals. Specific inclusion and exclusion criteria for the participants at the four levels are detailed in Table 2. The criteria have been developed through a Delphi-based expert consensus-building process.

**Table 2:** Criteria for participating in the program.

| Participant type | Inclusion Criteria | Rationale | Exclusion Criteria | Rationale |
| --- | --- | --- | --- | --- |
| Hospitals | 1. A Traditional Chinese Medicine (TCM) hospital or a hospital of integrated traditional Chinese and Western medicine at the second level or above;<br>2. Member of Guangdong Provincial Network of TCM Hospitals for the Management of Brain Conditions;<br>3. The hospital has a head CT that is open 24 hours × 7 days. | 1. TCM hospitals used Chinese medicine frequently in stroke management;<br>2. Using the network will ease the research implementation;<br>3. Head CT is necessary for the diagnosis of Stroke. | | |
| Departments | 1. Having no less than 20 beds reaches;<br>2. Stroke accounts for no less than 20% of annual inpatient admission;<br>3. The department head must be an attending physician or above and in stroke clinical work for more than 5 years. | Those three criteria ensure that we are targeting established stroke treatment providers. |  |  |
| Clinicians | Attending doctors or above who are managing stroke patients. | To ensure that the program reaches the intended clinicians. | Internship or rotation doctor. | To ensure that the program reaches the intended clinicians; |
|  |  |  |  | increase validity. |
| Patients | 1. Age $\geq 18$ years old ;<br>2. Diagnosed as non-cardiogenic stroke (Defined as 8B10,8B11,8B00,8B01,8B02,8B03 in ICD-11<sup>32</sup>), confirmed by head CT or head MR;<br>3. Hospitalized for stroke in a medical institution enrolled in this program.<br>4. The time of diagnosis is consistent with the time mentioned in the guidelines | 1. The guideline to be implemented is for adults;<br>2. The program targets patients with stroke;<br>3. The program targets inpatients.<br>4. Meet the applicability of the guidelines | 1. Diagnosed with pathological brain disorders, such as vascular malformations, tumors, and abscesses;<br>2. Diagnosed with other serious acute diseases including heart attack, asthma attack, pneumonia, appendicitis, organ failure, acute bronchitis;<br>3. Diagnosed as pregnant.<br>4. Repeat admissions | 1. Avoid delaying the treatment of other diseases.<br>2. Avoid delaying the treatment of other diseases.<br>3. Avoid the effect of this program.<br>4. Meet the applicability of the guidelines |

### 2.4 Sample size and sampling method

The program will target to recruit as many as possible from the 45 hospitals in the aforementioned TCM network. One patient admission seems as a sample. The sample size calculation is based on the primary implementation outcome, the physician’s adherence to the guidelines for stroke management concerning TCM, operationalized as a dichotomous variable describing whether to follow the stroke-management guideline or not (detailed later in 2.5.1).

In a complete factorial design, each factor will be allocated to half of the samples and the sample size depends on the smallest and clinically significant difference between the presence and absence of a factor. We power our study on the dichotomous variable “Adherence to the guideline-based stroke management”.

According to the formative work with clinicians at the TCM Hospitals in Guangdong Province, a relative risk of approximately 30% would be considered clinically significant. If <u>50%</u> of physicians(should meet the inclusion and exclusion criteria in 2.3) followed the guideline-based stroke management (estimates based on our prior work with clinicians), to detect a 30% difference(i.e., 65% of the samples followed the guideline). Assuming a two-tailed test, the alpha level of 0.05 and the desired power of 0.8, according to the formula below^33^, we can get the sample size of one experiment condition *N*_*srs condition*_ (*simple random sampling, srs*):

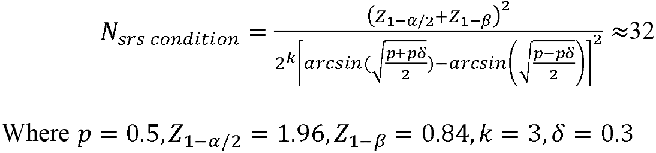

Therefore, approximately 256 samples are required to detect this effect (32 in each of the 8 conditions).

To avoid contamination, this study will consider conducting a between-clusters randomization approach, in which clusters (clinics or hospitals) are assigned as whole units for implementing experimental conditions. The clusters increase the variance of samples, which requires consideration of the design effect *D*_*e f f*_. We assume *D*_*e f f*_ = 2. Therefore, the final sample size of one condition (*N*_*complex*_) can be updated as follows:

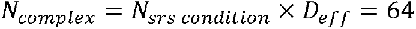

Therefore, 512 samples are required(64 in each of 8 conditions).

### 2.5 Data and data collection

#### 2.5.1 Outcomes

The outcomes to be measured have been determined through a modified Delphi expert process. The research team prepared the initial outcome sets, following the RE-AIM (reach, effectiveness, adoption, implementation, and maintenance) framework^34^. The initial measures domain of the effectiveness in the RE-AIM used the standard outcome set for stroke^35^, which was well developed by a group of internationally recognized experts (including one from China) from the International Consortium for Health Outcomes Measurement (ICHOM). The ICHOM standard outcome sets feature acute complications, disease control as well as a large component of patient-reported outcomes. The initial RE-AIM outcome sets went through two rounds of the anonymous consensus-building process. 10 experts participated in the Delphi process including 2 stroke-related physicians, 4 health system/service researchers, 3 behavioral scientists, and 1 health economist. The process led to the establishment of 24 outcomes within the five RE-AIM domains. All outcomes are deemed to be important to the stakeholders and also can be operationalized. The details of this process will be reported in a separate paper. The final outcome set, including the variable names, the definition of the outcome, the timeframe for collection, and the methods of collection, are summarized in Additional file 1. The primary outcome will be clinician adherence to guideline-based stroke management (Outcome 4.1.1, Additional file 1). The original guideline research team will develop this checklist. The checklist will include critical process indicators that represent the fidelity of the guideline implementation. The current draft checklist is listed in Additional file 2. The checklist will undergo further refinement. The primary outcome will be formulated according to the specific recommendations of the guidelines checklist, either as a continuous variable indicating the proportion (value 0 to 1) of what should be done in the checklist that is done or will be classified as a dichotomous variable (value 0 or 1), through which experts will judge whether the treatment process follows the guidelines or not. The information on the checklist completion will be abstracted from the medical chart.

#### 2.5.2 Implementation process data

We will conduct the process evaluation at the end of the program. Choosing this time point is to minimize the unintended influence on the project as the process evaluation itself may become a form of implementation technique if conducted during the program. We will use a method that we have developed and termed “**<u>S</u>**ilent & **<u>A</u>**nonymous **<u>FE</u>**edback (SAFE)” to obtain the process information. Our proposed method is inspired by the consensus-building techniques of the Delphi method and Nominal Group Techniques^36^. The purpose of the SAFE is to foster an equal-voiced rather than dominated discussion while removing participants’ fear of judgment or retribution during and after the meeting. Specifically, SAFE in this study will include the following processes in a 2-hour virtual group session: (1) preparing a list of categories of potential SAFE participants (such as administrators, physicians, nurses, financial personnel, support staff, etc) from an implementation site (i.e., the participating hospital), (2) from each category, asking the site to provide several potential participating names, (3) randomly selecting 1 person from the names under each category, (4) selected participants attending a web-based group meeting (such as using Tencent’s VooV Meeting) anonymously with a pseudonym, (5) at the virtual meeting, a facilitator opening the session to introduce the purpose and agenda, (6) the facilitator then releasing a web-based survey for the participants to fill out immediately, that contains a checklist of processes supposed to be completed during the guideline implementation, (7) survey results being immediately displayed on the shared screen with the participants, (8) the facilitator asking specific questions regarding why and how an item on the survey are performed from each participant’ perspective, (9) all participants being encouraged to write their response on the VooV meeting chat box that are visible to all participants and to respond to the postings of other participants if they would like to, and finally, (10) the facilitators asking any questions further if needed and then wrapping up. The survey involved in the process of SAFE will be further developed but will include the items about (1) how exactly the frameworks are applied, (2) challenges in applying the frameworks and the approaches adopted to meet those challenges, (3) specific guideline-implementation techniques developed as a result of applying the frameworks, (4) modifications made throughout the implementation to those techniques, and (5) participants’ rating of the use of the frameworks and the implementation techniques in facilitating the implementation. We should note this process is to solicit not only negative but also positive feedback. The format will give both quantitative (i.e., the survey results) and qualitative information (chat box conversation) for later analysis. We believe SAFE will be a more efficient way of engaging busy clinicians and hospital staff. The written response may also improve clarity in the expression of opinions and statements of facts. We will continue to develop more details of the process of SAFE and the survey forms to be used. We intend to publish a detailed protocol for doing SAFE on our project site (https://www.researchgate.net/project/ACACIA-Study) before we implement this in the field. In addition to the formal process of collecting process information through SAFE, we will ask the participating hospitals whether they have used any formative evaluation in their own implementation of the guideline. We would ask them to share the formative evaluation information (which may be qualitative or quantitative data) so that we can retrospectively analyze that data.

#### 2.5.3 Economic variables

When employing an implementation strategy, costs are incurred^37^. In the implementation research, there are mainly three costs: 1) Development and Execution of the implementation strategy; 2) Execution of the evidence-based practice; 3) Downstream costs including healthcare and non-healthcare resources caused by the intervention^38^. Implementation costs are mainly to develop and implement one or more evidence-based practices. In our study, as illustrated in Figure 1, we will include planning and preparation for the CFIR, TDF and NPT strategy, training of stakeholders and some changed behaviors related to the strategy. Intervention costs are resource costs directly from the consequence of implementation strategies targeting EBPs. It is recommended to distinguish the implementation costs and the intervention costs^39^. Apart from preparation and training of EBP, our study will also collect the changed resources needed to implement the intervention. Downstream costs are defined as those resources needed after finishing the intervention, including healthcare utilization and productivity costs of caregivers. In our study, we take the resources needed for audit and feedback, as well as assessing of maintenance as the downstream cost.

**Figure 1.**
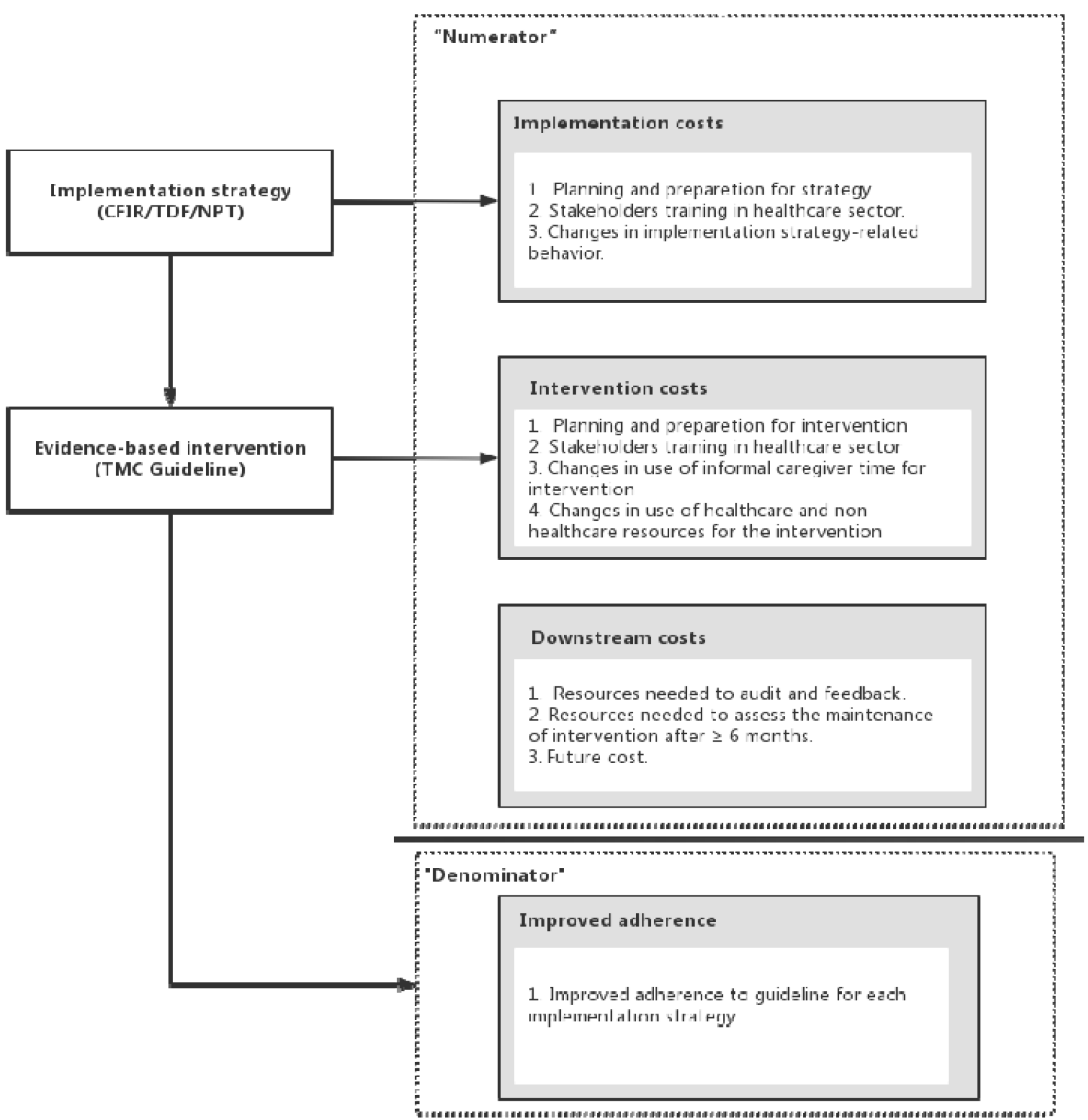
Types of costs to consider measuring and economic consequence (Modified from Heather T.Gold^38^)

To develop the cost estimates, we plan to use time-driven activity-based costing (TDABC)^40^. Firstly, we will calculate the capacity cost rate for the stakeholders (including clinicians, nurses, front workers, and managers of the healthcare sector, etc.) as a function of total annual compensation divided by the annual working time. Then, we will estimate the demand time of study-related behaviors according to Table 3. With the capacity cost rate and demand time, the costs for this study can be calculated.

**Table 3:**
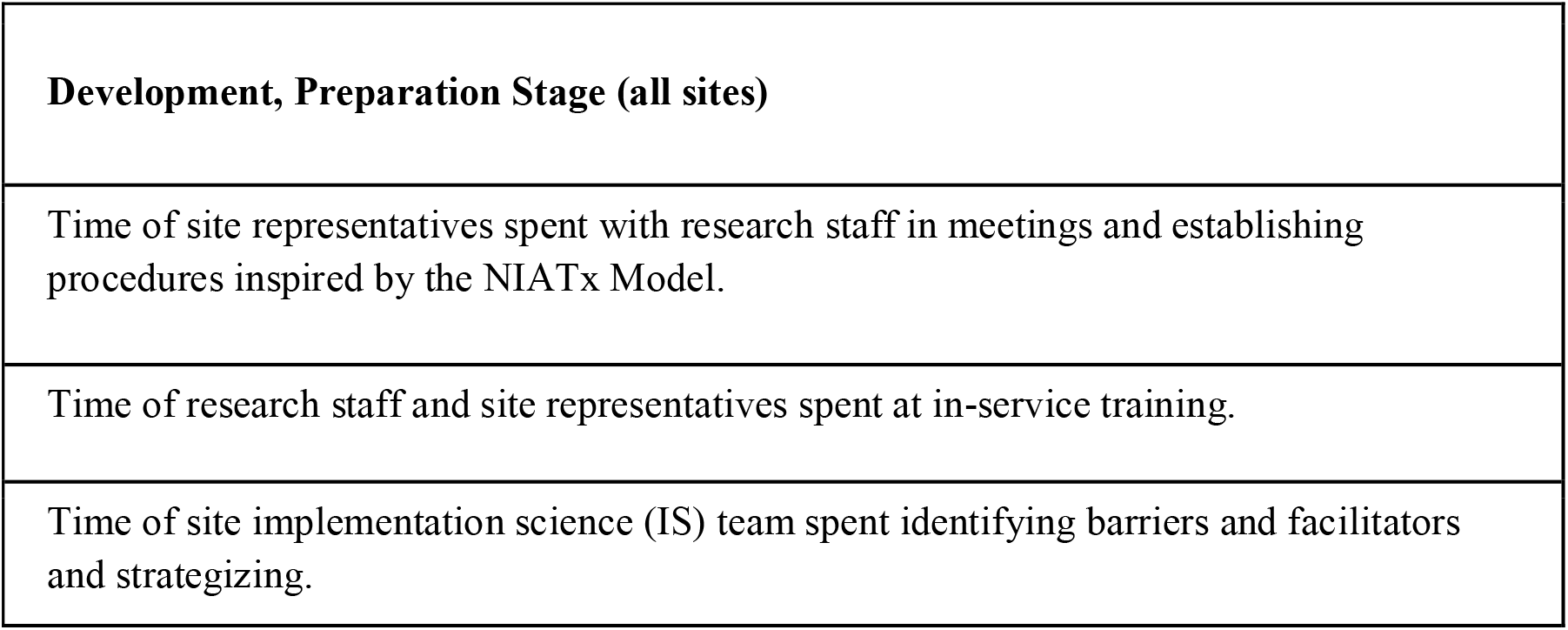
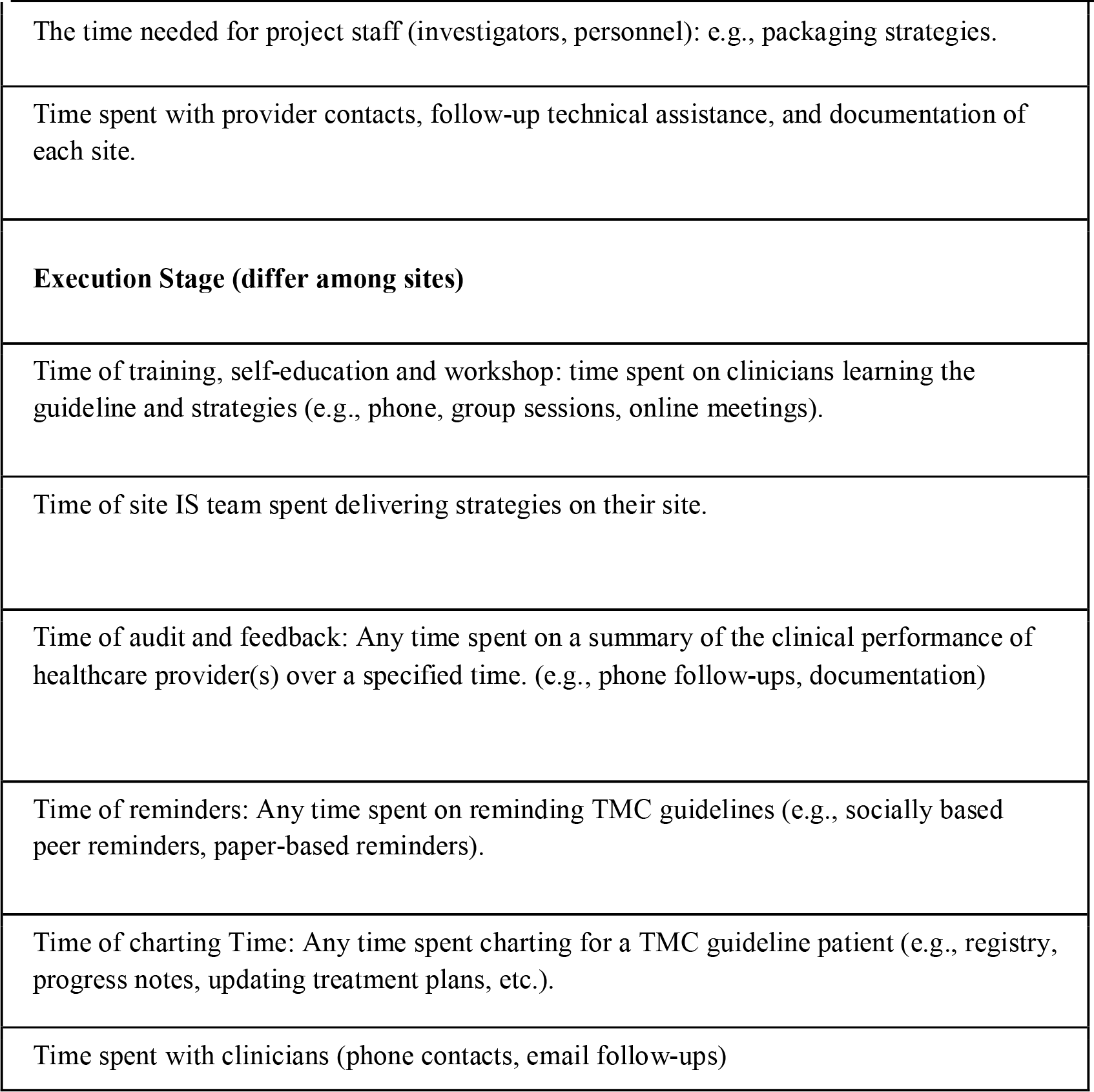
Recording time spent on the study (Implementation strategy, Intervention and Downstream behaviors)

### 2.6 Analysis

#### 2.6.1 Factorial analysis

Following an intent-to-treat model, a hierarchical logistic regression model will be used to test the hypotheses regarding the main effects of the three frameworks (TDF, CFIR, and NPT) and their interaction effects on the study’s primary outcomes. As described in the study design, this study is a cross-section of data with a three-level structure consisting of patients (level 1) nested within physicians (level 2) nested within hospitals (level 3).

##### Level-1 Model

Let*Y*_*i jk*_ be the outcome of patient *i* seeing physician *j* in hospital *k*. For a between-clusters experiment, one can model level-1 responses as

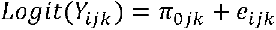

Where

*π*_0*jk*_ is the intercept for physician *j* in hospital *k*;

*e*_*ijk*_ is a level-1 random effect that represents a combination of random patient variability and measurement error. These residual patient effects are assumed normally distributed with a mean of 0 and variance *σ*^2^.

##### Level-2 Model

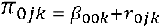

Where

*β*_00*k*_ is the intercept for physician in modeling the patient effect *π*_0*jk*_;

*π*_0*jk*_ is the level-2 physician random effect that represents the deviation of physician jk’s level-1 patients coefficient, *π*_0*jk*_, from its predicted value based on the physician-level model.

##### Level-3 Model

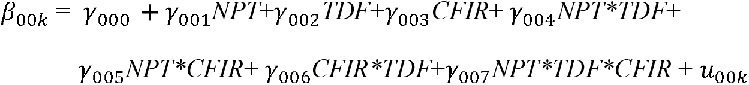

So that

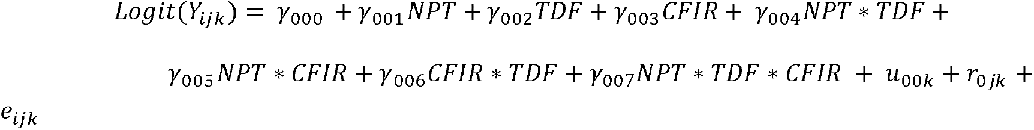

Where

*γ*_000_ is the intercept term in the hospital-level for *β*_00*k*_;

NPT, TDF, CFIR are three predictors for the hospital effect, including the main effect and two-way, three-way interaction effect. The predictor will be +1 if the hospital receives the experiment condition and -1 for the control condition^41^.

*γ*_001_ ∼ *γ*_007_ are the corresponding level-3 (cluster-level) coefficients that represent the direction and the strength of association of those three predictors’ main effect and interaction effect. *γ*_001_∼ *γ*_007_ expresses the half the expected change in *Y* when *NPT* or *TDF* or *CFIR* is taken from -1 to +1.

*u*_00*k*_ is a level-3 hospital random effect that represents the deviation of hospital k’s level-2 physician coefficient*β*_00*k*_.

The hierarchical logistic regression model not only can simultaneously assess the individual- and cluster-level variance but are also good at estimating the parameter when some data are missing using maximum likelihood if the missingness is Missing at Random (MAR) or Missing Completely at Random (MCAR)^42^. Further, we will examine all the data for missing information and loss to follow-up and will use multiple imputations as an alternative. Lastly, after well defining the hierarchical logistic regression model which best captures the design of the study, the model diagnostics will be performed to test the assumptions of the multilevel models.

#### 2.6.2 Implementation process analysis

The purpose of the process evaluation and analysis is to understand (1) what exact implementation techniques have been taken to implement the stroke guideline, and (2) how the frameworks have been used to guide the process of implementation. The quantitative information from the survey data (see section 2.5.2) will be tabulated descriptively with means, frequencies, and statistical tests when appropriate. The qualitative data will be analyzed with ATLAS.ti with Deductive Content Analysis (DCA)^43^. The deductive content analysis approach fits with our study as our analysis will be framework-guided. Specifically, two researchers with qualitative expertise will read the transcripts from the SAFE process afore-discussed and other sources repeatedly to become familiar with them; categorize important ideas and concepts in the material to a coding system informed by the implementation frameworks (specific frameworks to be decided later), and identify corresponding examples of excerpts from the material. The specific ways for each implementation team in applying the three frameworks understudy will be abstracted into a written summary. An expert panel with deep knowledge of those frameworks will evaluate the specific application of the frameworks for their appropriate use, misuse, and superficial use. All implementation techniques will be summarized and tabulated.

#### 2.6.3 Economic analysis

We plan to perform Cost Effectiveness Analysis (CEA)^44^ to figure out whether it will be worth adding cost considering the benefits of each component. The economical outcome variables include (1) improved adherence at different component combinations, and (2) the cost per percentage of adherence saved. Incremental cost-effectiveness ratios (ICERS)^45^ will be used to compare the cost and benefit of three implementation framework components. The ICERS will be defined as ΔC/ΔE, ΔC is the cost a component adds while ΔE is the effectiveness (ΔC and ΔE are the numerator and denominator respectively in Figure 1). Traditionally, quality-adjusted life year (QALY) is an effective factor to measure effectiveness. However, we can not get a quantitative relationship between adherence to the guideline and the QALY in stroke patients. Therefore, adherence will be directly used for effectiveness. So, ICERS means what the cost will be if the increase of one percentage of adherence to the guideline. In some cases, if 1 intervention costs more than another, it still is a good choice if it improves adherence at an acceptable cost. The cutoff of the acceptable cost will reach a consensus by SAFE. To get a robust selection of the components, we will also estimate the 95% confidence interval around the ICERs using bootstrapping simulation methods^44^.

#### 2.6.4 QCA analysis

To investigate which combinations of outer setting features in conjunction with the use of different frameworks may act as necessary or sufficient conditions for the occurrence of the outcome (successful implementation of stroke clinical guidelines), we propose to use qualitative comparative analysis (QCA). Based on Boolean logic^46^, QCA compares sets (a set that can be defined as a group of elements that share certain characteristics) of conditions and the relationship of conditions to outcomes by examining across-case patterns. QCA’s examination of cross-case patterns acknowledges the diversity of cases and their heterogeneity with regard to their different relevant conditions (“variables”) and contexts by comparing cases as configurations. Rather than assuming random data distribution, QCA is a non-linear and non-additive method investigating how observations are distributed across rows in a “truth table” (a data metric with 2^k^ rows, where k is the number of conditions; the truth table reflects all logically possible combinations of conditions, with each row refers to a specific combination of conditions (i.e., a configuration)). Applying the Quine-McCluskey algorithm (method of prime implicants), the truth table reflecting all logically possible configurations can be reduced to Boolean equations that minimize these combinations which yield prime configurations. QCA uses two measures to assess goodness-of-fit: consistency (i.e., the strength of the link between condition to outcomes, within the range of 0-1) and coverage (i.e., the fraction of cases to which relationship applies, within the range of 0-1) to assess goodness-of-fit. The QCA analysis will be performed using the R software (R Core Team 2020, Rstudio text editor 2020) and the R packages “QCA”^47^ and “SetMethods”^48^.

We will use an explanatory sequential design^49^ to collect qualitative data with key stakeholders from a small number of representative hospitals to help inform the quantitative and qualitative data collection with the full study sample. We will select three high- and three low-performing hospitals from the full sample of participating hospitals for qualitative data collection using two criteria. First, we evaluate the baseline performance of participating hospitals by using the RE-AIM domains (as discussed in more detail in the earlier section) and rank the hospitals on their baseline performance. Second, we purposively sampled hospitals representing diversities of geographic location, size, and ownership.

We will employ one-to-one and focus group interviews with key stakeholders who involve directly or play a central role in implementing stroke management clinical guidelines, including hospital senior or middle-level managers, hospital administrators, patient care managers, clinicians, patients, patients’ family members or care supporters, etc. The qualitative data will be analyzed by using ATLAS.ti (qualitative analytical software) to inform the selection of the most relevant setting features in conjunction with the use of frameworks in implementing stroke management clinical guidelines in the study. Using the QCA method, we treat each participating hospital as a case (the unit of analysis), given the case-oriented nature of interest in the present research.

### 2.7 Discussion

In this proposed factorial cluster RCT, we will examine the main effect and interaction of the use of implementation frameworks in facilitating the implementation of the stroke management guideline in traditional Chinese medicine Hospitals in the Guangdong province of China. Qualitative comparative analysis will be used to analyze the contextual factors that facilitate or hamper the implementation of the guideline. The study will be a type III hybrid design that focuses on testing the effect of the implementation strategies (the use of the implementation frameworks) while collecting and analyzing the effect of the health and clinical outcomes.

As of November 17, 2022, we have not found other studies that study the effectiveness of using implementation frameworks in facilitating the implementation of a clinical intervention using an experimental design. As implementation research is rapidly developing, we need to develop empirical evidence on the strengths and weaknesses of using implementation frameworks. Furthermore, the study can provide experiences on how implementation strategies can be tailored to the context of the organization. In the trial for the effectiveness of a clinical intervention, consistently conducting clinical intervention across organizations is pursued. However, for implementation techniques, many a time they will have to be context-specific. In this trial, although each organization will be applying the implementation frameworks in a standardized way, they have the flexibility of producing different implementation techniques and packaging those into implementation strategies (bundle of those implementation techniques) that suit the outer and inner contexts of their organization. In addition to the examination of applying implementation frameworks, the method of QCA will enable us to examine the contextual factors. QCA has been used more retrospectively in the past, which has the limitation of having no information on some conditions of interest. In this trial, we will be able to use theories and empirical experiences to help us prospectively plan for the data collection. Finally, the process evaluation of this study will also yield useful information for a comparative understanding of different frameworks and their application in real practice. Recommendations will also be made on how those frameworks might be applied for future studies considering their strengths and weaknesses.

There are several foreseeable limitations of the study. First, the facilitators and the hospital implementation teams are not blinded to the frameworks to be applied to their work. However, unlike a biomedical trial, we expect the placebo effect of this implementation trial to be minimal. Second, it is possible that the hospitals may tap into ideas from other implementation frameworks not assigned to them in their actual implementation work. Our choice of cluster randomization may help reduce this possible contamination.

## Supporting information

RE-AIM indicator system for the implementation of stroke guideline.

Checklist of guideline implementation

Standards for Reporting Implementation Studies: the StaRI checklist for completion.

## Data Availability

All data produced in the present study are available upon reasonable request to the authors

## Declarations

### Ethics approval and consent to participate

This study was approved by the Southern Medical University review board (IRB; #202261). Informed consent will be obtained from all participants for surveys and qualitative interviews. All methods were carried out in accordance with relevant guidelines and regulations. Research involving human participants, human material, or human data, must have been performed in accordance with the Declaration of Helsinki.

### Consent for publication

Not applicable.

### Availability of data and materials

The data taht support the findings of this study are available from the corresponding author upon reasonable request.

### Competing interests

No competing interests

### Funding

Z Chen acknowledges financial support from the General Program of the National Natural Science Foundation of China (Grant No. 72174098)

### Author’s contribution

DR Xu, YF Cai, and YY Cai designed the study. DR Xu and W He wrote the manuscript. CH, ZC, and YS provided suggestions and improved the health economics evaluation section. PF Guo provided guidance on factorial design. S Lv, LP Zhang, and Q Zhao provided guidance and suggestions on expert consensus. L Liu provided guidance and supplementation on QCA methodology application. All authors read and approved the final manuscript.

## Acknowledgements

We would like to express our sincere gratitude to the teachers from the Guangdong Provincial Hospital of Traditional Chinese Medicine for their valuable contributions to this project.

## Notes

### Competing Interest Statement

The authors have declared no competing interest.

### Author Declarations

Ethics committee/IRB of Southern Medical University gave ethical approval for this work

