## Supplementary material for "Effectiveness of using implementation frameworks to facilitate the implementation of a stroke management guideline in the Traditional Chinese Medicine hospitals in China: protocol for a factorial randomized controlled trial": RE-AIM indicator system for the implementation of stroke guideline.

### Additional file 1. RE-AIM indicator system for the implementation of stroke guideline.

| **Indicators** | **Calculation** | **Collection Time** | **Instruments/Source** |
| --- | --- | --- | --- |
| **1. Reach：number, proportion and representativeness of patient participants** | | | |
| 1.1 Number of patients consenting to participate in the project | Number of patients who met the program recruitment criteria and signed informed consent to participate in the program | Baseline survey | Informed consent signed by patients when they joined the program |
| 1.2 Proportion of the number of participating patients in the target population of the project | Molecule: the number of patients who meet the project patient recruitment criteria and agree to join the project  Denominator: The number of patients who meet the program patient recruitment criteria at the program participating institution | Project patient recruitment period | Molecule: informed consent signed by the patient when he or she joined the program  Denominator: hospital records of stroke patients |
| 1.3 Representativeness of patients participating in the project  1.3.1 Representativeness of patients recruited to the program | The case-mix of recruited patients and eligible patients was compared at baseline.  The case mix consisted of a set of variables associated with stroke outcome. | Baseline survey | ICHOM standard set of stroke outcome measures |
| 1.3 Representativeness of patients participating in the project  1.3.2 The representativeness of patients who participated in the entire program | At the end of the program, the case mix of patients who had participated throughout the program and those recruited at baseline was similar | Project the whole process | ICHOM standard set of stroke outcome measures |
| 1.4 Percentage of patients who drop out of the program | Molecular: Number of patients who quit the program early before the program end point  Denominator: total number of patients enrolled in the program  At the end point of the project, the case-mix of patients who participated in the project and those who dropped out were compared | Project the whole process | Molecular: Project participant's missing records  Denominator: informed consent for patient participation in the project |
| **2.Effectiveness：The effectiveness of the project at the patient level** | | | |
| 2.1 Main clinical indicators：smRSqscore-Ability to return to  usual activities | See remarks for scoring rules | Baseline, discharge, 7 days after discharge, 90 days after admission (i.e., end point) | Clinical specialists interviewed patients or their families using the smRSq scale |
| 2.2 Secondary clinical indicators：All-cause mortality | Patient dies (Yes/No) | 90 days after discharge and admission | Medical records and patient follow-up questionnaire |
| 2.3 Secondary clinical indicators: recurrence rate | Self-reported new  stroke (Yes/No) | 90 days after admission | Patient follow-up questionnaire |
| 2.4 Patient-reported（PROMIS-10）  2.4.1 Smoking cessation | Smoking(Yes/No) | 90 days after admission | Have you ever smoked since you were hospitalized for stroke |
| 2.4 Patient-reported（PROMIS-10）  2.4.2 Functional status after stroke | Mobility、Self care (including grooming, toileting & dressing)、Feeding、Ability to communicate | Discharge, 7 days after discharge, 90 days after admission | ICHOM questionnaire entries（PROMIS-10) |
| 2.4 Patient-reported（PROMIS-10）  2.4.3 Overall physical  wellbeing (including  pain, fatigue, and  general health status) | General health, general quality of life, general physical health, general mental health, general social activity and relationship satisfaction, general ease of daily activities, general mood (anxiety, depression), and general pain in the past 7 days | Discharge, 7 days after discharge, 90 days after admission | ICHOM questionnaire entries（PROMIS-10) |
| **3.Adoption：The number, proportion and representation of institutions and physicians involved in the project** | | | |
| 3.1 Organizational indicators  3.1.1 Proportion of participating institutions | Molecule: the number of institutions that meet the recruitment criteria and agree to join the program  Denominator: Number of institutions that meet the recruitment criteria of the program | Project agency recruitment period | Molecule: informed consent signed by the institution when it joined the project  Denominator: Medical institution questionnaire |
| 3.1 Organizational indicators  3.1.2 Representativeness of participating institutions | Similarity between participating institutions and all institutions eligible for recruitment (see remarks for variables used for comparison) | Baseline survey | Medical institution questionnaire |
| 3.1 Organizational indicators  3.1.3 Withdrawal rate of participating institutions | Numerator: The number of organizations that quit the project early before the end of the project  Denominator: The number of all institutions involved in the project | Project the whole process | Molecular: Institution withdrawal letter  Denominator: Prior to the start of the study, the participating institutions signed a consent form |
| 3.1 Organizational indicators  3.1.4 Representative of institutions participating in the whole process | Similarity between institutions that dropped out of the study and all institutions that participated at baseline (see remarks for variables used for comparison) | Project the whole process | Medical institution questionnaire |
| 3.2 Indicators of doctors  3.2.1 Absolute number of doctors participating in the project | Numerator: number of eligible physicians who have agreed to join the program  Denominator: Number of doctors eligible for recruitment | Baseline survey | Molecule: the consent form signed by the physician when he joins the program  Denominator: Medical institution questionnaire |
| 3.2 Indicators of doctors  3.2.2 Proportion of doctors participating in the project | Molecule: number of physicians who meet the program recruitment criteria and agree to join the program  Denominator: Number of physicians that meet the program's recruitment criteria | Baseline survey | Molecule: informed consent signed by doctors when they join the program  Denominator: Medical institution questionnaire |
| 3.2 Indicators of doctors  3.2.3 Representativeness of doctors participating in the project | Similarity between participating physicians and all physicians eligible for recruitment (see remarks for variables used for comparison) | Baseline survey | Doctor basic information questionnaire |
| 3.2 Indicators of doctors  3.2.4 Withdrawal rate of participating doctors | Molecular: Number of physicians who quit the program early before the end of the program  Denominator: Total number of doctors or nurses participating in the program at baseline | Project the whole process | Molecule: Notification of doctor's withdrawal from the program  Denominator: consent form signed by participating physicians before the study began |
| 3.2 Indicators of doctors  3.2.5 Representativeness of doctors participating in the whole project | The similarity between the doctors who participated in the program and all the doctors who met the recruitment criteria | Project the whole process | Molecule: Notification of doctor's withdrawal from the program  Denominator: consent form signed by participating physicians before the study began |
| **4.Implementation：The extent to which the guidelines are administered, local adjustments to their implementation, and the costs of their implementation** | | | |
| 4.1 Indicators of doctors  4.1.1 Adherence to the guideline-based stroke management | Follow the guideline (Yes/No) | hospitalization | Evaluated by the panelist according to the record from the hospitalization system. |
| 4.2 Organizational Level  4.2.1 Fidelity of institutional implementation Guidelines | Percentage of stroke clinical practice guidelines that completed key items during the study period | hospitalization | hospitalization |
| 4.2 Organizational Level  4.2.2 Institutional cost of intervention | Monetary cost to institutions implementing stroke intervention guidelines during the study period | Discharge and End points | Patient discharge cost list, patient and physician cost questionnaire |
| **5.Maintenance： Maintenance of the project after the study** | | | |
| 5.1 The degree to which stroke guidelines are normalized | Normalizing Value of NPT Scale | Endpoint (when the project ends as a study) | NPT scale |
| 5.2 Fidelity of intervention guidelines for institutional implementation after ≥6 months | For new patients 6 months after program completion: Percentage of institutions implementing stroke clinical practice guidelines | 6 months after the study ended | A random sampling of cases |
