## Supplementary material for "Effectiveness of using implementation frameworks to facilitate the implementation of a stroke management guideline in the Traditional Chinese Medicine hospitals in China: protocol for a factorial randomized controlled trial": Checklist of guideline implementation

### Additional file 2. Checklist of guideline implementation

| **No.** | **Compliance Items** | **Guideline Items** | **Observed** | | **Remarks** |
| --- | --- | --- | --- | --- | --- |
|  |  |  | **Yes** | **No** |  |
| **Ischemic stroke** | | | |  | |
| **1.** | **Patients:**  Received medical treatment within 6 h after onset | **3** |  |  |  |
|  | **Treatments:**  Include Buyanghuanwu decoction and Xuefuzhuyu decoction when using rt-PA. Or include Notogiseng Radix Et Rhizoma. when using urokinase |  |  |  |  |
| **2.** | **Patients:**  Not received medical treatment in the first 5 h after onset | **3** |  |  |  |
|  | **Treatment:**  Not recommended to use Chinese herbal medicine that activates blood and resolves stasis. |  |  |  |  |
| **3.** | **Patients:**  Patients with ischemic stroke | **2** |  |  |  |
|  | **Treatment：**  Chinese herbal medicine can be used in a complementary manner for secondary prevention of ischemic stroke to reduce the risk of a recurrence,    disability, and mortality in the long term. Specific clinical characteristics, personal preference, and financial status should be considered on a case-by case-basis when deciding whether to prescribe Chinese herbal medicine. (2C) Chinese herbal medicines with potential benefits include Dengzhanshengmai capsules, which are often used in addition to conventional strategies for the secondary prevention of ischemic stroke. |  |  |  |  |
| **4.** | **Patients:**  Patients with ischemic stroke who are to ameliorate neurological impairment | **1** |  |  |  |
|  | **Treatment:**  Chinese herbal medicine can be used in addition to conventional care and treatment to ameliorate neurological impairment in patients with ischemic stroke. Chinese herbal medicines that activate blood and resolve stasis are widely used. Greater benefit is likely when Chinese herbal medicine is started in the acute stage and continued for 3-30 days. (2C) Chinese herbal medicines with potential benefits include Dengzhanxixin injection, Shenxiong glucose injection, Buyanghuanwu decoction, injections made from Salviae Miltiorrhizae Radix Et Rhizoma, and proprietary Chinese medicine made from Notogiseng Radix Et Rhizoma, injections made from Ginkgo Folium, and herbal formulae that act as purgatives. |  |  |  |  |
| **Hemorrhagic stroke with coma** | | | | | |
| **1.** | **Patients:**  With hemorrhagic stroke with  coma | **6** |  |  |  |
|  | **Treatments:**  Xingnaojing injection could be used as a complementary treatment to improve the level of consciousness |  |  |  |  |
| **2.** | **Patients:**  With poststroke coma, which is defined as a syndrome of internal blockage of phlegm-heat in Chinese medicine. | **6** |  |  |  |
|  | **Treatments:**  Angong Niuhuang pills could be used as an adjunct to conventional care to improve the level of consciousness.The duration of treatment is usually 10-14 days. |  |  |  |  |
| **Transient ischemic attack** | | | | | |
| **1.** | **Patients:**  Who have experienced a transient ischemic attack with no signs of macroangiopathy | **2** |  |  |  |
|  | **Treatment:**  Chinese herbal medicine could be an alternative to pharmacotherapy to reduce the risk of recurrence and the possibility of developing an ischemic stroke. |  |  |  |  |
| **2.** | **Patients:**  Patients have experienced a transient ischemic attack | **2** |  |  |  |
|  | **Treatment:**  Chinese herbal medicine can also be used in addition to a conventional secondary prevention strategy for a greater reduction in the risk of recurrence or an ischemic stroke. |  |  |  |  |
| **Ischemic stroke with hemorrhagic transformation** | | | | | |
| **1.** | **Patients:**  Ischemic stroke with hemorrhagic transformation | **4** |  |  |  |
|  | **Treatments:**  Chinese herbal medicine that activates blood and resolves stasis could be used in addition to conventional care and treatment after hemorrhage is stabilized, to reduce neurological impairment |  |  |  |  |
| **Hypertensive intracranial hemorrhage** | | | | | |
| **1.** | **Patients:**  Not candidates for surgery. | **5** |  |  |  |
|  | **Treatments:**  Chinese herbal medicine that activates blood and resolves stasis is safe when used in addition to conventional care and treatment. The contraindications should be considered carefully before treatment, and any clinical decision regarding the specific timing for initiation of Chinese herbal medicine should take the enlargement of the hematoma and the risk of recurrent hemorrhage into account. Chinese herbal medicines with potential benefits include Xueshuantong injection and Naoxueshu oral solution. |  |  |  |  |
| **2.** | **Patients:**  With surgically treated | **5** |  |  |  |
|  | **Treatments:**  In view of its beneficial effects on mortality, hematoma, and neurological impairment, Chinese herbal medicine that activates blood and resolves stasis can be used in addition to conventional care and treatment in patients with surgically treated hypertensive intracranial hemorrhage. The contraindications should be reviewed closely before starting treatment with Chinese herbal medicine, and the clinical decision making regarding when to start it should take into account whether or not there is an enlarging hematoma and the risk of recurrence of hemorrhage. |  |  |  |  |
| **Dysphasia after stroke** | | | | | |
| **1.** | **Patients:**  Patients with dysphasia after stroke. | **7** |  |  |  |
|  | **Treatments:**  Acupuncture can be used in addition to routine care and rehabilitation, including swallowing training. Acupuncture prescriptions with potential benefits include needling at GV26 (Chinese name: Shuigou), GB20 (Fengchi), EX-HN12 (Jinjin), EX-HN13 (Yuye), and CV23 (Lianquan), which is usually for 4 weeks. |  |  |  |  |
| **2.** | **Patients:**  Patients with dysphasia after stroke. | **7** |  |  |  |
|  | **Treatments:**  Chinese herbal medicine could be used as a complementary strategy to improve swallowing in patients with dysphasia after stroke. (2C) External application of Chinese herbal medicine, such as stimulation with herbal ice-sticks or sprays, has potential benefits. |  |  |  |  |
| **Poststroke depression** | | | | | |
| **1.** | **Patients:**  Patients with poststroke depression. | **8** |  |  |  |
|  | **Treatments:**  Chinese herbal medicine that soothes the liver and regulates qi could be used in addition to antidepressants for additional improvement of depressive signs and symptoms in patients with poststroke depression. (2C) Chinese herbal medicines with potential benefits include Chaihushugan powder, Xiaoyao powder, Shuganjieyu decoction, and Shuganjieyu capsules. |  |  |  |  |
